# Interprofessional falls prevention in community settings: Older adults’ perspectives through a socioecological lens

**DOI:** 10.64898/2026.07.12.26357305

**Authors:** Amy Lawton, Rebecca Wospil, Nicholas Tripodi, Danielle Baxter, Brett Vaughan, Professor Rebecca Lane, Jack Feehan

## Abstract

**Objectives:** Falls among community-dwelling older adults are a global public health concern. Although falls prevention care relies on collaboration between medical and allied health professionals, older adults’ experiences and perspectives of interprofessional care remain underexplored. This research therefore explored the perspectives and experiences of primary falls prevention care, in order to identify what care is occurring in practice, where preventative opportunities may be missed and how older adults would prefer falls-prevention care to be delivered.

**Design:** Qualitative exploratory study with a pragmatic orientation and utilising focus groups for data collection. Data was analysed using reflexive thematic analysis, with themes developed iteratively and subsequently interpreted through the lens of the Socioecological Model to develop a consumer-informed socioecological framework for community falls prevention care.

**Setting:** Three metropolitan (Melbourne, Adelaide, Brisbane) and two regional centres (Woodend, Wagga Wagga) in Australia.

**Participants:** Thirty-six older adults participated across five focus groups.

**Results:** Four themes were identified: knowledge of allied health and falls prevention, access to care, co-ordination of care, and when and how to provide care. Each theme reflected interacting influences across socioecological levels, demonstrating the complexity of older adults’ engagement in falls prevention.

**Conclusions:** Despite strong support for preventative care, participants’ experiences were frequently consumer-driven, reactive and fragmented. Limited knowledge of falls and allied health, unclear access pathways, and inadequate coordination of care, shaped by organisational and policy contexts, were identified as key barriers. System-level reform that embeds falls prevention and allied health within routine care and aligns funding, coordination mechanisms and public health strategies to deliver equitable, sustainable prevention are required.

**Strengths and limitations of this study:** • This qualitative focus group study provides novel consumer insight into older adults’ experiences of allied health engagement in interprofessional community-based falls prevention care.

• The application of a socioecological framework enabled interpretation of falls prevention care across individual, interpersonal, community, organisational and broader system levels.

• Inclusion of participants from diverse health service contexts enhances the transferability of findings, however, the findings may not be generalisable to all contexts and may reflect the views of older adults who were sufficiently engaged in healthcare to participate.

## Background and Objectives

Falls among community-dwelling older adults remain a major global public health concern as a leading cause of injury and death (World Health Organization, 2021). Approximately one-third of community-dwelling older adults will fall each year, increasing to two-thirds in those with a history of prior fall or fear of falling (Salari et al., 2022). Other consequences of older adults experiencing a fall are significant and include ongoing physical disability, reduced quality of life, persistent fear of falling, loss of confidence in physical abilities, social isolation, increased caregiver dependency, and early admission into aged care (Hopewell et al., 2018). With an increasing life expectancy globally, the ageing population will require increased access to acceptable and equitable falls prevention services (Montero-Odasso et al., 2022). Despite strong evidence supporting primary falls prevention activities, implementation within routine community care remains inconsistent and fragmented (Delbaere et al., 2025).

Current falls prevention guidelines advocate a multifactorial approach to screening, assessment and intervention, alongside the need for interprofessional collaboration (IPC) (Montero-Odasso et al., 2022). However, evidence shows that adoption of fall prevention guidelines by health professionals varies widely and does not reliably translate into changes in practice or reduced falls, highlighting persistent implementation gaps in real-world settings (Haley et al., 2025). Interprofessional collaboration is defined as multiple healthcare professionals (HCPs) from different disciplines providing collaborative, coordinated care (Interprofessional Education Collaborative, 2023). As a range of disciplines distinct from medicine or nursing (Lewis et al., 2025), allied health (AH) plays an integral role in IPC for falls prevention through screening, assessment and intervention across physical, environmental, psychosocial and behavioural domains (Montero-Odasso et al., 2022). As a collective, AH professionals account for approximately one-third of the global health workforce, with their numbers increasing at a faster rate than the medical and nursing professions (Lewis et al., 2025). Despite their growing numbers and broad clinical scope, current guidelines do not represent the breadth of AH professions who can contribute to falls prevention care. In the face of global healthcare workforce constraints (World Health Organization, 2022), alongside evolving aged care standards and practices, it is essential that the scope of all AH professions is understood and effectively utilised to provide quality preventative care.

Individual engagement with AH and interprofessional falls prevention care is shaped not only by service availability, but also by how care is understood, accessed, coordinated and delivered (Vincenzo et al., 2022). Implementation of multifactorial falls prevention in community settings is influenced by health professional behaviours, organisational support, and consumer contextual factors,, affecting how and when services are used (Vandervelde et al., 2023). Effective IPC in community settings depends on provider communication, shared processes, and coordination of care. The Socioecological Model (SEM) provides a framework examining how individual, interpersonal, organisational and broader system factors collectively shape engagement with falls prevention care (McLeroy et al., 1988).

Despite the complexities of providing primary falls prevention care, older adults’ perspectives, including their understanding of AH roles, navigation of care pathways, and experiences of co-ordination are underrepresented in the primary falls prevention literature (Montero-Odasso et al., 2022). Importantly, understanding primary falls prevention requires attention to the perspectives of those who have not fallen or experienced care previously. Older adults who have not fallen may have different expectations of prevention, preferred ways of accessing care, and views on the roles of health professionals because their perspectives have not yet been shaped by a falls or secondary falls prevention care. Therefore examining these perspectives can help identify what is currently occurring in practice, how older adults understand the contributions of different AH professionals, where opportunities for prevention may be missed and how older adults would prefer to access and receive this care.

This study therefore aimed to explore Australian older adults’ perspectives on community-based falls prevention and IPC, including their understanding of AH roles and experiences of accessing and navigating care. The study included people who had and had not experienced a fall and falls prevention care to examine perceptions of healthcare professional roles, identify desire and opportunity for primary prevention and understand how they would prefer this care to be delivered. Using the Socioecological Model (SEM) as an interpretive lens (McLeroy et al., 1988), the study sought to identify how individual, interpersonal, organisational and broader system factors shape these experiences. These insights may inform strategies to meet unmet healthcare needs that older adults experience duet to limitations in the availability, accessibility, affordability and acceptability of services (Rahman et al., 2022),

## Qualitative Research Design and Methods

Ethical approval was received from *name redacted for peer review* University (HRE approval code redacted for peer review). This study is reported in accordance with the Consolidated Criteria for Reporting Qualitative Research (COREQ) guideline (Tong et al., 2007).

## Study design

An exploratory qualitative study with a pragmatic orientation was undertaken to explore older adults’ experiences and perspectives on interprofessional community-based falls prevention. Focus groups were utilised for data collection to facilitate discussion of shared and differing experiences of falls prevention care, with group interaction enabling participants to compare and build on each other’s perspectives(Stewart & Shamdasani, 2014).

## Participants and Recruitment

Potential participants were English-speaking, community-dwelling adults over 60 years of age and included those with or without a history of falls or prior falls prevention care in order to capture those approaching older age and primary prevention perspectives. There were no exclusions due to sex, gender or cultural background. Participants were purposively recruited by the authors through a mixture of face-to-face and online contact, via the research teams’ professional networks, community groups, online consumer health forums, and social media advertising. Potential participants were invited to contact the lead author via phone, email or online form using the Qualtrics platform (Silver Lake, USA), to express interest in participating. The lead author then contacted interested participants via their preferred contact method to supply verbal or written study information as appropriate to the method.

## Data Collection

A semi-structured focus group topic guide was developed (Stewart & Shamdasani, 2014) and piloted with two older adults who were not involved in the study following ethics approval (Supplementary material A). No revisions were made to the guide following piloting or during data collection. The focus group topic guide explored participants’ perceptions and experiences of community falls prevention care including AH roles, collaborative care, and triggers for accessing primary falls prevention care.

All participants provided written informed consent prior to completing the demographic survey and participating in a focus group. Demographic data and other data related to individual participant falls prevention was collected via paper or online survey using the Qualtrics platform (Silver Lake, USA). Consent for audio-recording was verbally re- confirmed at the beginning of the focus group, before recording commenced. A list of the AH professionals included in the survey and focus group discussions was informed by Allied Health Professions Australia (Allied Health Professions Australia, n.d.). The list was provided to each participant before completing the demographic survey and focus group, and time was provided to review the list and ask any clarifying questions of the researchers.

Focus groups were held in person at accessible community venues local to participants between April and October 2025 and lasted approximately one hour. At the beginning of the focus groups, the motivations for the research, researchers’ professional backgrounds and roles, and the study aims were explained to participants at the beginning of each focus group. Lead author (AL) facilitated the focus groups, complementing the semi-structured guide with open-ended prompts and follow up questions to clarify responses, elicit examples and explore participants’ experiences and perceptions in greater depth. Participants were encouraged to compare and contrast their experiences to others in the group, and the facilitator invited less vocal participants to contribute and checked for differing perspectives before moving to a new topic.

A second member of the research team (RW, NT or BV) attended to support session logistics, document contextual observations and field notes, and contribute to a post-session reflexive discussion with the facilitator to support the consideration of researcher influence, group dynamics and preliminary analytic impressions, No other non-participants present, and no prior relationship existed between the researchers and participants.

## Data Analysis

Focus groups were audio recorded and transcribed using Microsoft 365 software (Microsoft Corporation, USA), then checked and corrected by the lead author for accuracy and removal of identifiable information. Transcripts were not returned to participants for verification or feedback.

Inductive reflexive thematic analysis (RTA) was employed to identify and interpret patterns and shared meanings related to the research objectives, with a focus on real-world experiences, barriers, enablers and opportunities to improve care(Braun & Clarke, 2019, 2021a). Analysis followed Braun and Clarke’s six-phase approach to RTA, moving iteratively between familiarisation, coding, theme generation and review. Researchers AL, RW and NT engaged with the transcripts repeatedly, recording initial observations and reflections before undertaking inductive coding in NVivo (Lumivero, USA). Ongoing discussion and reflexive engagement with the data were used to explore different interpretations and challenge assumptions. Candidate themes were developed by bringing together related codes around broader concepts and revised against the coded extracts and the wider dataset. Themes were named and defined according to their central concepts and relationship to research questions. Focus group locations were classified based on the Modified Monash Model (MMM), an Australian Government classification system that categorises locations from metropolitan (MM1) to very remote (MM7) based on remoteness and population size (Versace et al., 2021). These classifications were considered during the data analysis to explore whether patterns in participants’ experiences and perspectives differed across location categories.

Consistent with RTA, data saturation was not sought; instead, the adequacy of the analysis was determined by the depth and richness of the interpretation in relation to the research aims (Braun & Clarke, 2021b).

## Framework development

Following completion of the thematic analysis, authors BV and AL identified that the described across the themes aligned with the levels of the SEM. The themes were therefore subsequently interpreted through the SEM as a second analytical step.

Authors AL and RW mapped coded data within each theme across individual, interpersonal, community, organisational, and systems/policy levels in NVivo. Framework development then involved identifying modifiable influences within each theme and SEM level and considering how these presented opportunities to improve interprofessional falls prevention care. A draft framework was reviewed by all authors who provided feedback on the interpretation and alignment of the findings and recommendations. The framework was iteratively refined until consensus between authors on structure and content was reached.

## Author reflexivity

All research team members are AH professionals and tertiary educators with experience in qualitative research methods. The researchers’ professional backgrounds and interest in gerontology, falls prevention and health systems helped shape the research focus and interpretation. The lead researcher (AL) and co-authors (RW and NT) who analysed the data are osteopaths with over 13 years’ experience in community-based care for older adults. Their professional backgrounds in osteopathy may have influenced the emphasis on AH roles in the analysis.. Reflexivity was supported throughout using field notes completed after each interview which documented observations and preliminary interpretations and regular meetings during data analysis to discuss themes, researcher biases, and potential alternative interpretations.

Patients or the public were not involved in the design, or conduct, or reporting, or dissemination plans of our research.

## Results

### Participants

Thirty-six (n=36) older adults aged 66–89 years participated in five focus groups. Fifty-six individuals initially registered interest in the study, of whom 38 were able to attend one of the scheduled focus groups. Two participants subsequently withdrew on the day of their focus group due to illness. A summary of the participants’ demographic data can be found in Table 1.

**Table 1:**
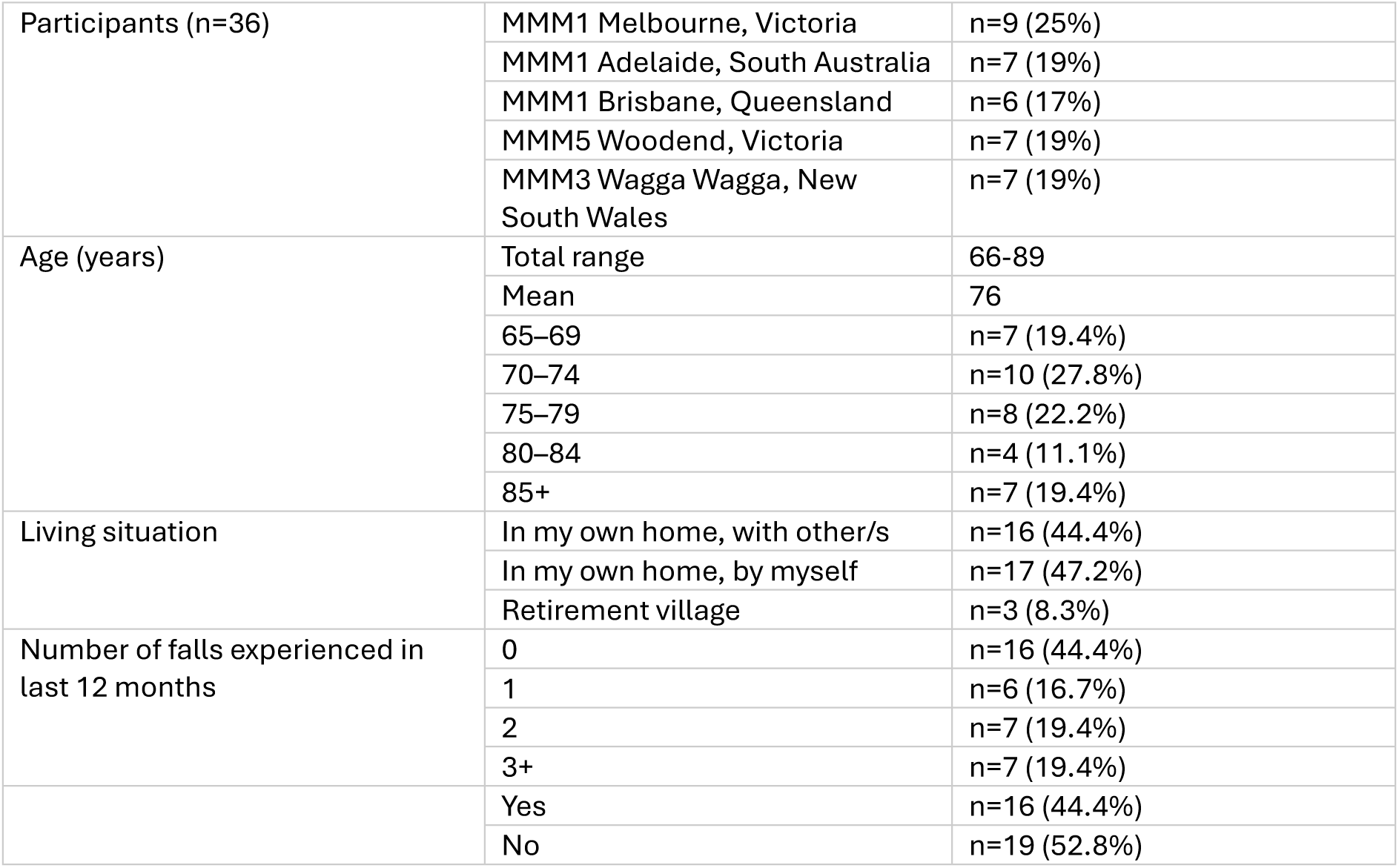
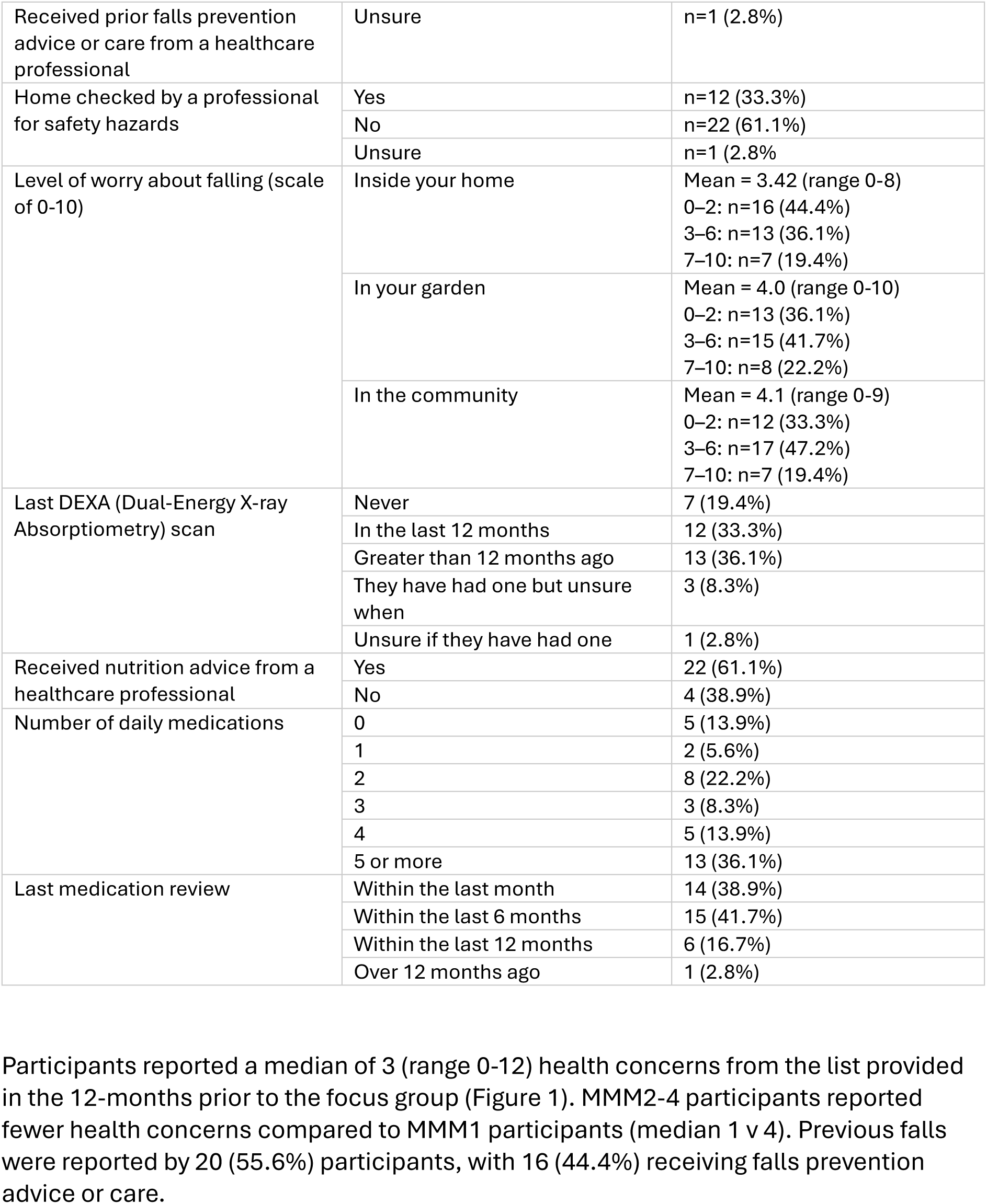
Consumer focus group participant demographics.

Participants reported a median of 3 (range 0-12) health concerns from the list provided in the 12-months prior to the focus group (Figure 1). MMM2-4 participants reported fewer health concerns compared to MMM1 participants (median 1 v 4). Previous falls were reported by 20 (55.6%) participants, with 16 (44.4%) receiving falls prevention advice or care.

**Figure 1.**
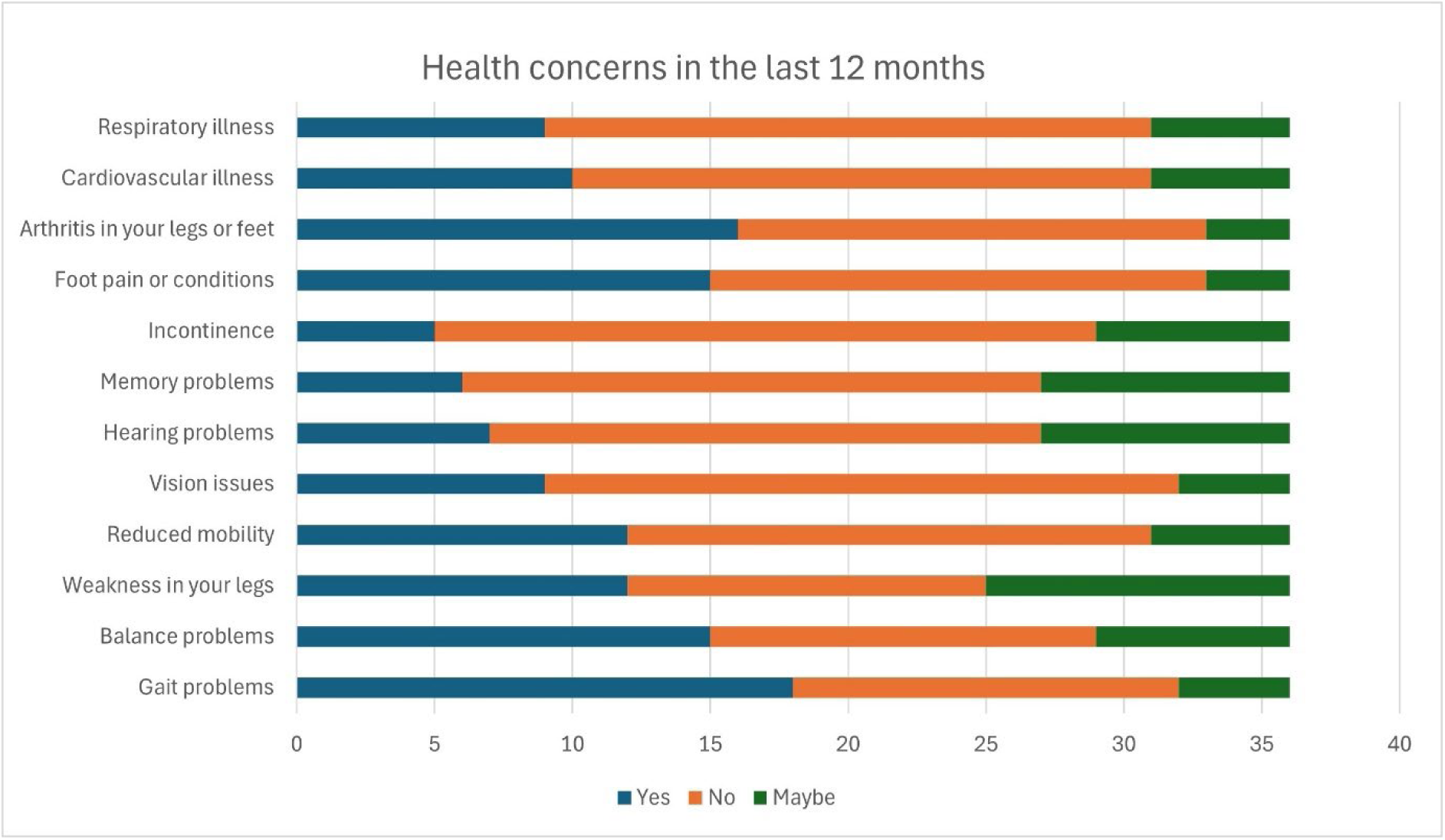
Participant health concerns in the previous 12-month period. Figure created by the authors using Microsoft Excel Alt text Figure 1 Horizontal stacked bar chart of 12 health concerns experienced in the previous 12-month period

Participants who reported receiving prior falls prevention care indicated this was provided by a physiotherapist (PT) (n=7), occupational therapist (OT) (n=2), doctor or nurse (n=5), with one participant being referred to a falls clinic. All participants reported previous engagement with at least one AH professional in the 12 months prior to the focus group. Participants accessed a median of 7 AHPs (range 1-15), with MMM1 participants reporting an average of 8 interactions, compared to 6.5 for MMM2-4 participants (Figure 2). Pharmacy was the most consulted AH profession, followed by physiotherapy and podiatry, with progressively lower levels of previous contact reported across other professions (Figure 1). No participants reported previous contact with genetic counselling or music therapy.

**Figure 2.**
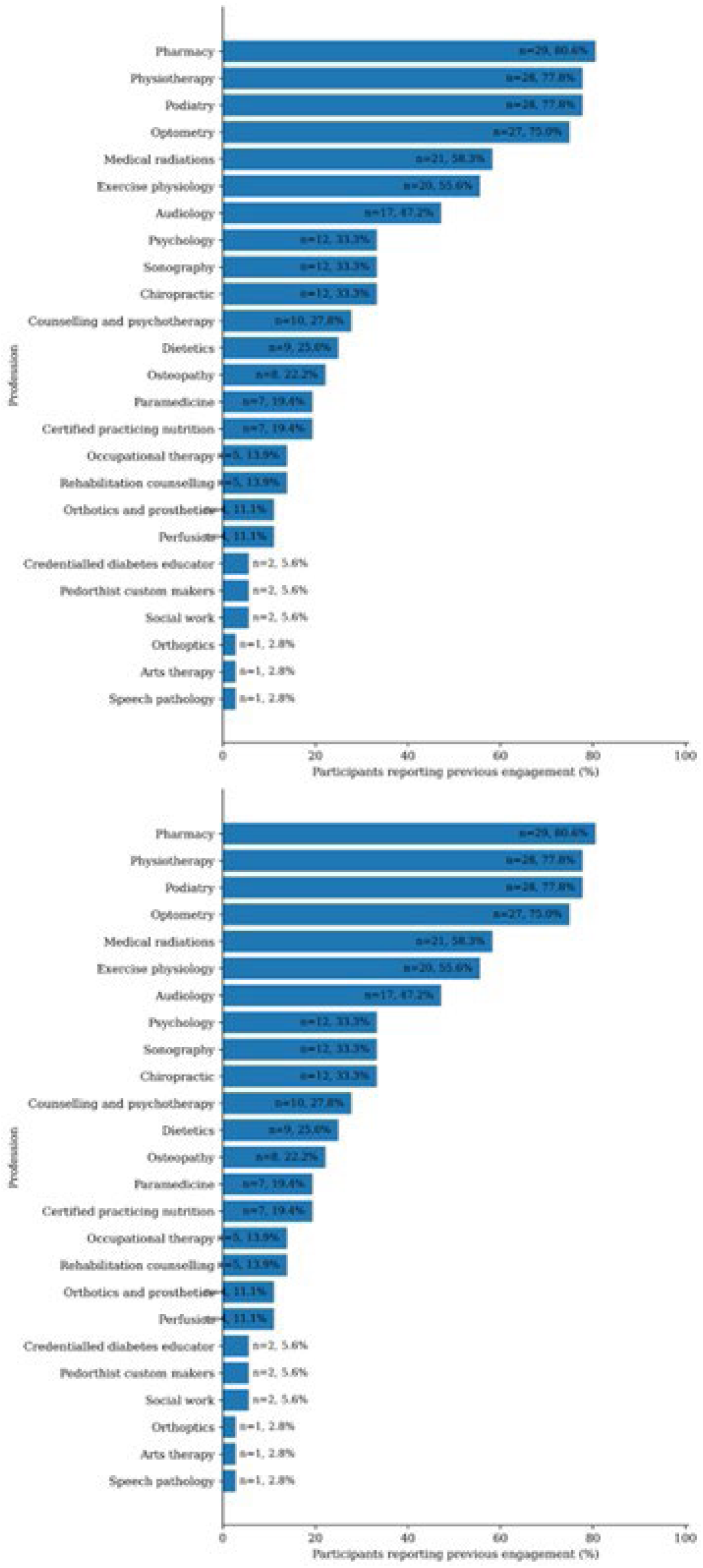
Previous engagement with allied health professions among participants, shown as number of respondents (n) and percentage (%). Figure created by the authors using Microsoft Excel Alt text Figure 2 Previous engagement with allied health professions among participants, shown as number of respondents (n) and percentage (%)

### Thematic analysis

Reflexive thematic analysis generated four themes reflecting how participants experience and view AH and IPC in community falls prevention care: knowledge of AH; access to care; co-ordination of care; and when and how to provide care (Table 2).

**Table 2:** Themes.

| **Theme** | **Descriptor** |
| --- | --- |
| ***Knowledge of falls prevention and AH*** | Captures participants' understanding of falls prevention and allied health within the healthcare system. It includes awareness, or lack thereof, of who are considered allied health professions, what they do, how their roles relate to falls prevention. It also includes influences of uncertainty, misconceptions and gaps in knowledge and system literacy. |
| ***Access to care*** | Describes participants' experiences of accessing allied health services and falls prevention care. It includes referral pathways, professional gatekeepers, funding and appropriate service availability. It also reflects logistical, financial and system-level factors that enable or constrain entry into care. |
| ***Co-ordination of care*** | Represents participants' perspectives on how care is connected or fragmented between individual HCP or between services. It includes communication between providers, information sharing, referral processes and responsibility for co-ordinating care. It also captures the burden placed on older adults when they are required to organise, sequence or communicate their own care. |
| ***When and how to provide care*** | Reflects participants' views on the appropriate timing, context and models of delivering falls prevention support. It includes reflections on when primary preventative care should begin, adequacy of funding models, opportunistic and routine health checks, repeated messaging and community-based programs. It also encompasses the participants' desire for a public health approach, including awareness campaigns that can normalise prevention and increase reach. |

**Table 3:** SEM Levels.

| **SEM Level** | **Descriptor** |
| --- | --- |
| Individual | Encompasses personal factors such as knowledge, beliefs, attitudes, skills and behaviours that influence health decisions. |
| Interpersonal | Reflects the influence of relationships with family, friends, caregivers and healthcare providers on health behaviours and decision-making |
| Community | Considers the broader social environments in which people live, including community resources and services that shape opportunities for engagement and social participation. |
| Organisational | Relates to the structures and processes within healthcare services that influence knowledge, access and the delivery of care. |
| Policy & enabling environment | Encompasses system-level factors such as health policy, funding models and regulatory frameworks that impact health service access and delivery. |

## Knowledge of allied health and falls prevention

At the Individual level of the SEM, participants routinely reported a lack of understanding of who AH professionals are and what they do, not only in the context of falls prevention, but across healthcare more broadly.

> *What do you mean by allied health? Like who you’re referring to?* (FG5, P6)

> *There seems to be a lot of different areas that people work. Are they under that banner or under a main banner …this music therapist, occupational therapist anything with therapists? Are they all individual?* (FG2, P5)

Interactions with HCPs were viewed as key drivers of falls and AH knowledge, with the general practitioner (GP) identified as the key enabler for developing this knowledge but also a potential barrier.

> *…with both my falls, my doctor* [GP] *has checked me out, but nothing else has happened. And it was because of that that I’ve sort of worked out my own way of looking at the things that caused the fall.* (FG3, P9)

> *It would be interesting to know how each of these people* [medical professionals] *feel about the other* [allied health] *professionals.* (FG2, P7)

Non-health community organisations were seen as key resources for improving falls and AH knowledge, in what is viewed as a public health issue.

> *Because, you know, they [community organisations] have a mandate around public health, this [falls] is a public health issue. And I think that they should be, you know, doing more in that in this space They’ve got a lot of opportunity to be doing things…it’s about health prevention and prevention, you know, it needs to come from a whole different range of different areas. (FG4, P3)*

> *I reckon that it’s such a major thing that there should be a preliminary place that you can go to, to get advice to keep yourself informed as to the steps to take in your life so that the risk of falling is minimised. (FG3, P9)*

Organisational models which include co-location of medical and allied health services and shared medical records were perceived to aid knowledge acquisition, as well as access to and coordination of care.

> *With an EPC [Enhanced Primary Care plan] again there, the allied health person is supposed to send back a report and so that ties your GP in. But if you’re fortunate to have two allied health in the one building,which I do, then they liaise, but only on my instigation…if you can find some way to get them to work in well that would be great. (FG2, P2)*

Participants emphasised that older adults need clear guidance on which HCP to approach for falls prevention care. This is aligned with participants’ views that investing time and funding in preventative care is both desired and necessary to improve long-term system costs.

> *Most people who, a lot of the people don’t know who to go to. Too late. They’ve had a fall. It costs the government a lot of money to look after them whereas if they had information beforehand… (FG3, P9)*

> *If they don’t have time to look at you and get that information, you’re going to be back in hospital and costing the system a lot more. I just, you know, prevention is the thing, but it comes to cos*t. (FG4, P9)

## Access to care

Participants felt that it was their responsibility to initiate falls prevention care and identified the need to advocate for themselves when visiting their general practitioner (GP) to enable access to appropriate referrals.

> *I mean, how are they gonna [sic] know that you’re worried about a fall unless you actually bring it into the conversation? (FG1, P9)*

> *He’s [GP] very good at giving referrals, but I have to be the one to suggest, well, who I get referred to, for sure. (FG3, P9)*

Access to, or information about, funding was reported as an important role of the GP. Funding meant that the participants could access the care that they perceived they required without significant impact on their personal finances. However, some participants reported hesitancy from their GPs to refer to AH.

> *When you get older, like I don’t know about everyone here, but money does play a part in what you can and can’t do. (FG5, P5)*

> *…GPs don’t seem to want to send you to something like osteo [osteopath] or physios [physiotherapy] or acupuncture’ (FG1, P3)*

Finding a trusted AH professional with the knowledge and skills that are needed for the individual was also discussed as a key enabler to access acceptable care. Regional participants found that limited services in their area could result in them travelling further distances to access appropriate care.

> *But what is it? Do you kiss a lot of frogs before you find a Prince?* (FG4, P5)

> *It’s also a very general sort of term about what they* [allied health professionals] *actually do. They deal with kids, they deal with elderly…so you need a specialist OT other than just a generalist OT.* (FG3, P10)

Community-led services were viewed by participants as an opportunity to scale care, provided the local community is aware of their availability and how to access them.

> *…getting one-on-one is gonna* [sic] *be difficult as we’ve all realised that maybe these sort of group sessions through different organisations U3A* [University of the 3^rd^ Age]*, Men’s shed, ladies shed, you know things like that that you might be able to get a group of people together that so that they can then find out what, virtually a bigger, bigger thing than what we’ve got here. So that would allow more people to have access.* (FG3, P4)

> *…there’s a huge need for the awareness of those services and I suspect that they don’t advertise a lot because their funding comes and goes…if there’s more consistent funding, maybe they’re more likely to advertise their services.* (FG2, P7)

Differences across hospital, community and private healthcare organisations impacted participants’ ease of access, particularly through existing variation in structured referral pathways and communication between HCPs.

> *So that’s an interesting way of building up medical practices. So, they’re clearly doing that in the hospital system but perhaps there needs to be more of that. (FG2, P10)*

Participants viewed funding as an enabler to access AH care but also demonstrated limited understanding of where AH sits in the healthcare system and how to access their services.

> *I’ve recently gone through My Aged Care to get an appointment with an exercise physiologist, hopefully to do the falls and balance clinic. (FG2, P2)*

> *I thought, yeah, the GP could only do that part of the referral. (FG5, P4)*

## Co-ordination of care

Participants discussed the need to be proactive when it came to coordinating their own care, rather than relying on their GP or other health professionals to do so.

> *We can’t blame everyone [health professionals]. We’ve gotta look after results. We’ve gotta have that information. So that’s all there is to it. We’ve gotta be educated more. (FG4, P9)*

Logistically, coordination of healthcare created a challenge for the participants where they reported needing to time their care from each health professional and share their health records between the health professionals to facilitate improved care.

> *…so it’s pointless going to see the first one* [health professional] *unless you’ve been to see the second one* [health professional] *because they don’t have the information. So you gotta get them all your ducks lined up. And it’s very hard to say, well, I’m going to see X here. Then I wanna see Y. Then I wanna see Z then and then the whole three of them. Those results will go back to A and then a can make an overall assessment. OK, this is what’s come back out of all the results.* (FG4, P8)

Participants described experiencing limited coordination of care. They viewed GPs and practice nurses as the primary HCP who should be co-ordinating care, due to the referral rights and funding access required to fulfil this role.

> *GPs are the catalyst, they refer off, but once that referral’s gone, they don’t know whether or they’ve given you a referral. They [GP] don’t know whether you followed up on that referral or not. (FG2, P9)*

Organisational processes such as regular health checks were identified by the participants as a strategy to increase confidence that their care will be well managed and coordinated. Conversely, operational factors such as appointment length and practitioner workload were seen to limit preventative care.

> *I feel really confident in my [healthcare] practice, that every six months someone’s gonna [sic] know everything about my health. Yeah, and they’re gonna [sic] steer me in the right direction. (FG3, P7)*

> *And they’re [GPs] all very busy, right . You know, they’ve got enough to do within what’s happened in the session with you and then, you know, bringing up other matters, their times limited. (FG1, P7)*

The appropriateness of GPs as the central point of contact was discussed by participants, with several advocating strongly for a GP-centric system. However, most participants supported prioritising access and skillset over professional title.

> *It mightn’t be the GP who is so very, very busy. Maybe there’s like medical coordinators in places whose role is to do the coordination and the education*.

> *Maybe you’ve got a different role because I mean, GPs are very, very busy people*. (FG2, P7)

## When and how to provide care

Participants indicated a strong desire for preventative care, and this appeared to be a key driver for seeking out their GP to create referrals to other health professionals.

> *I’d rather have information before if all happens, right, so that I know to alter my my footwear, to not have rugs in my house, to know not to fall over my dog. (FG4, P3)*

> *So, if you can stop that first fall, they’ll, you know, live and keep living a nice and healthy life. (FG5, P8)*

Access to personal medical records was consistently viewed as supporting understanding and agency of care.

> *In terms of in terms of self help that there is room for guidance like falls and risks of falling in relation to every consumer’s access to their own their own medical records. (FG1, P4)*

Falls prevention care was seen as the responsibility of all HCPs by the participants. However, a culture of practice which encourages preventative care was identified as being required for participants to build trust in the system, with some participants associating this with an allied health model of care.

> *I just think being informed constantly about that* [falls risk] *is really important*. *Whoever you go to*. (FG2, P3)

> *…I don’t believe that they* [medical specialists] *will actively direct you to ways of managing your life, so you don’t fall. I think it’s allied health way above rheumatologists, neurologists, they’re interested in the disease but not how to work around it.* (FG2, P2)

Community organisations were identified by the participants as an opportunity for social interaction and peer learning, with the prospect of repeated messaging. In regional areas, this was enhanced for participants who are aware of and access community transport services.

> Hm*m, well, I think that’s where your community neighbourhood houses come in. Because you bring in the social, you know, loneliness is another thing…it doesn’t have to be a lengthy…activity that you do around falls prevention provided it’s done at a lot of different opportunities… (FG4, P3)*

> *You mentioned about the groups getting together but getting together as a group. Just information from each other. (FG3, P8)*

> *Well, the* [location] and [location] *neighbourhood houses have a community transport programme that you can use and is subsidised, but yeah, that’s again an awareness thing… (FG2, P2)*

Participants reflected on increasing awareness and the need for advertising of falls prevention programs run by local organisations to increase participation.

> *These are things you need to be aware of or are you aware that the local Community Health Centre has a falls and balance programme? Maybe go and talk to people there. (FG2, P9)*

> *I’m wondering if advertising has got anything to do with this because coming back to (location name redacted) after being away for so long, I have no idea where to go and I had no idea where to settle in. I didn’t understand anything of it and so if there was some way that there was advertising out there so we knew where you go. (FG3, P5)*

The opportunity to incorporate allied health expertise into a local hub that provides falls workshops was consistently posited as an acceptable way to provide care.

> *But it would be nice to be able to incorporate their experience and expertise into a hub. Yeah, where they could give advice at a different time. (FG3, P7)*

> *I would like to see more help in sort of like a a workshop and how to, when you do fall, you know, what do you do then? You know, I’ve seen people in a situation where they don’t know they can’t get up. And yeah, there are ways and I know there are ways. I think it’s it’d be good to have that run by the allied health or something like that? (FG4, P7).*

### Policy/enabling environment

An age-appropriate approach to equitably increasing population-level awareness of falls and services through age-appropriate channels is desired. This could be through public health campaign or triggered by existing processes such as driver’s license renewal or existing yearly health checks.

> *Think it’s a bit of a problem with advertising in our current world in the younger people coming through are just doing it on social media and things like that. And like you, don’t have a computer. There’s a lot of people in the community who aren’t hearing about things because they’re not using the old ways of advertising as well. There needs to be much more campaign and advertising to reach everybody because everybody deserves to get the information. (FG3, P7)*

> *There’s an opportunity there to be proactive and actually send something out with a with a pension advisor email or something that says look, we’re going to have an occupational therapist coming out into your region this date. (FG4, P8).*

Participants repeatedly emphasised that systems and funding mechanism design need to support individuals in accessing care through multiple health and community pathways to increase reach. They also recognise that these systems need to enable organisations to sustainably advertise and provide their services through consistent funding to improve awareness and engagement.

> *When you get older, like I don’t know about everyone here, but money does play a part in what you can and can’t do. And I think that there should be more avenues available for the older people to exercise and do that sort of stuff. Because a lot of the falls, if you had the strength, you would prevent yourself from falling, I think. (FG1, P5)*

> *And there’s a huge need for the awareness of those services and I suspect that they don’t advertise a lot because their funding comes and goes. So it’s, you know, So if there’s more consistent funding maybe they’re more likely to advertise their services. (FG1, P7)*

## Discussion

This study explored older adults’ perspectives of community-based allied health and interprofessional falls prevention care. Participants described care as reactive and fragmented with responsibility frequently shifting to the individual to recognise risk, initiate care, and navigate services. Limited visibility of AH roles, reliance on GP-centred funding and referral models, and inconsistent co-ordination shaped experiences of care. Despite this, participants expressed a clear preference for preventative care including both AH and medical professions, containing exercise, home modification, nutrition and addressing fear of falling, as reflected in current guidelines (Montero-Odasso et al., 2022).

### Individual level influences

Engagement with AH and falls prevention care was shaped by older adults’ knowledge, beliefs, experiences, and capacities. Participants in the current work reported having the responsibility to initiate conversations about falls concerns, which misaligns with current guidelines recommending that these conversations be initiated by HCPs (Montero-Odasso et al., 2022). This reliance on self-initiation is problematic in the face of potential embarrassment and fear of losing independence (Wiseman et al., 2025), relying on an individual’s trust in HCP relationships, rather than a system-level approach to preventative care. Although greater consumer engagement in care, including initiation, can improve treatment outcomes and patient satisfaction (Marzban et al., 2022), healthcare consumers are not consistently aware of what preventative falls care entails and how it relates to routine care (Mazza et al., 2011). If access to care is not provided, participants may self-manage perceived falls risk, or the lack of tangible action by the HCP may delegitimise preventative actions (Mazza et al., 2011).

Once the need for falls prevention care was established, participants described a responsibility for advocating for allied health referrals and coordinating their own care, despite limited knowledge of falls prevention and AH roles. The systems knowledge required for advocacy and navigation is produced through experience, formal knowledge and the capacity to undertake research, alongside economic and social capital (Willis et al., 2016). Capacity to engage in these processes can be constrained by co-morbidities and cognitive challenges, reliance on informal support networks, affordability and funding, systems and technological literacy, and access to clinical records (Dawkins et al., 2021; Vincenzo et al., 2022; Willis et al., 2016).

System navigation is further complicated by inconsistency in scope of practice definitions, AH profession overlaps in skillset, and variation of services offered by practitioners from the same profession (Downie et al., 2023). Although, exercise-based professions, particularly physiotherapy and exercise physiology, and occupational therapy were most frequently associated with falls prevention, they represent AH professions with established difficulties in overlapping skillset and role visibility (Rahja & Laver, 2019; Toloui-Wallace et al., 2024). With prompting, participants were able to retrospectively identify other AH falls prevention roles, for example optometry for problems with vision, which highlights the complex interplay between knowledge of falls and allied health to aid system navigation (Willis et al., 2016). Further research is required to understand why these AH professions were not readily identified in this care context, alongside exploration of AH and medical HCP perspectives.

### Interpersonal level influences

Access to falls and related health knowledge among older adults was largely experiential, with GPs consistently positioned as gatekeepers to care and coordination due to their control of referrals, funding, and high consumer trust. Falls prevention is viewed as the responsibility of all HCPs (Montero-Odasso et al., 2022), however, participants rarely experienced primary falls prevention practices, particularly when past falls were not voluntarily disclosed. Proactive information seeking by older adults is challenging (Mc Grath et al., 2016), and participants identified a trusted health professional as essential to accessing falls care, however even when presenting to a HCP after a fall, participants were rarely asked about contributing factors or referred to AHPs for further care. These missed opportunities have the potential to shape perceptions of health care (Dawkins et al., 2021), potentially delegitimising preventative care provided by AHPs (Mazza et al., 2011). The experiences described may partly reflect limited practitioner knowledge of AH roles, which can act as a barrier to referral and interprofessional collaboration (Grant et al., 2015; Liddle et al., 2020)

Participants expressed a strong desire for coordinated, person-centred falls care that considers interpersonal circumstances and integrates flexible, low-burden approaches into existing healthcare touchpoints. Opportunistic screening, brief education, and consistent messaging are all key to person-centred falls care (Eastman et al., 2022).

Whilst participants trusted GPs to coordinate care, they were open to AH or dedicated coordinators fulfilling this role, provided they possessed requisite knowledge and skills. This opening presents an opportunity for improved AH workforce utilisation that requires development of referral pathways, funding models, and accessible clinical records (Khong et al., 2017; Mazza et al., 2011; Dawkins et al., 2021).

Falls care should consider caregiver responsibilities and social support, prioritising flexible, low-burden approaches integrated into existing healthcare and community touchpoints to provide repeat messaging across multiple encounters. Care delivery should prioritise respect, empathy, and adequate time for listening to foster motivation and engagement with prevention strategies, enabling healthcare professionals to empower older adults through health education and supported decision-making (Khong et al., 2017). Future research could explore the barriers that prevent GPs from routinely assessing falls risk and referring older adults to AH services following a fall.

### Community level influences

Community level influences were less frequently discussed by participants compared to other SEM levels potentially reflecting limited visibility and integration of support services, rather than a lack of relevance. Whilst community organisations were identified as potential resources for falls and AH education, exercise interventions, and self-management support, access was perceived to be constrained by limited awareness, poor integration with health services and inadequate resources. Although there is evidence that community service providers are supportive of falls prevention, limited resources and confidence in providing associated activities are a barrier to falls care provision (Markle-Reid et al., 2015).

Participants identified trusted, accessible, age-appropriate community settings and resources as opportunities for falls prevention information, peer learning and social connection to support long-term engagement (Khong et al., 2017; Naseri et al., 2025). Commonly represented opportunities include community noticeboards or newspapers, HCP running community workshops through Neighbourhood Houses, Men’s Sheds or 3UA, gyms and leisure centres, and council expositions (Khong et al., 2017). Further, participants proposed a well-advertised community hub for falls prevention information housed within one of these organisations to increase confidence in accessing help in order to mitigate fragmentation between services. Future research should explore with consumers how falls prevention information and messaging should be shaped to effectively inform them of community-based falls care opportunities.

### Organisational level influences

Organisational influences on falls prevention care were most evident in the contrasts between private and community care settings, reflecting their differing structures, processes and policies which impact provision and coordination of care. Integrated care models featuring co-location of professions, appointment availability, shared clinical software, and established referral pathways can enhance access and coordination (Goodwin, 2016; Levesque et al., 2013). Community health organisations were viewed as particularly valuable in providing these features, and affordable care, though access varied geography, with fewer services in regional areas reflecting global challenges in healthcare accessibility and equity (World Health Organization, 2022).

In contrast, private healthcare organisations lacked structured organisational processes such as predetermined referral pathways and communication procedures, contributing to fragmentation where services operate across independently without shared coordination mechanisms (Auschra, 2018; Goodwin, 2016).

Hospitals, universities, and larger non-government and not-for-profit organisations such as Council on the Ageing (COTA) Australia were identified as valuable intermediaries in addressing organisational gaps through population-level reach and awareness promotion, particularly for regional older adults (Zhong et al., 2023). Effective falls prevention likely depends on organisational structures that support multidisciplinary teams through horizontal service integration, vertically coordinated care pathways across primary and community settings, and integrated networks linking programs within and across sectors. Collectively, these findings show that effective falls prevention depends on organisational structures that support multidisciplinary teams through horizontal service integration, vertically coordinated care pathways across primary and community settings, and integrated networks that link programs within and across sectors (Auschra, 2018; Goodwin, 2016). Future research should investigate strategies to better integrate private allied health practitioners into structured falls care pathways and the broader healthcare system to improve access to quality coordinated care and patient outcomes.

### Policy/enabling environment level influences

At the policy and enabling environment level, participants identified gaps in systems literacy, funding, and cross-system communications as constraints to falls prevention, emphasising the need for a whole-system approach integrating public health strategies with person-centred care to provide equitable, accessible falls prevention care (World Health Organisation, 2021a). A coordinated public health campaign to improve falls, allied health, and systems literacy for healthcare professionals and older adults is needed, as health literacy, perception, and trust are key barriers to preventative care engagement (Cross-Barnet et al., 2019).

Funding policy emerged as a central enabler and constraint for the participants in the current work. Whilst My Aged Care and Medicare were viewed as key enablers, limitations included reinforcing reliance on time-poor GPs, limited visit numbers under some of the funding schemes, and health workforce shortages. These findings are consistent with evidence showing contemporary funding models reinforce siloed practice and inadequately support healthcare professionals and consumers to engage in falls prevention (Grant et al., 2015; Liddle et al., 2020).

Blended funding models and policies supporting referral rights for non-medical professionals are needed to promote inclusion of AH, interprofessional collaboration and coordination capacity. Existing age-related health touchpoints and administrative processes such as driver’s license renewal present opportunities for consistent screening and early intervention. As many people may not engage with services such as government pensions and aged care funding until after the age of 65, it should be considered which touchpoints can reach the largest population for timely primary prevention. Further, improved access to shared national health records requires consideration of digital literacy and privacy concerns amongst older adults (Cross- Barnet et al., 2019).

If system and policy-level improvements expand access to services, local service availability must be addressed to meet increased demand, with strategies including student and community-led services, telehealth, non-profit organisations, and improved AH access through community health organisations contributing to a whole-systems approach (Gizaw et al., 2022). Future research should explore the effectiveness of blended funding models and potentially non-medical coordinator roles in supporting IPC for falls prevention at scale, particularly in regional areas.

### Consumer-informed socioecological framework for community interprofessional falls prevention care

The findings demonstrate that older adults’ experiences of falls prevention care are shaped by interacting influences across multiple levels of the healthcare system. Knowledge of AH, access to care, co-ordination, and perspectives on when and how to prevention should be delivered were influenced by individual factors, as well as healthcare relationships, service organisations, and broader policy and funding structures.

To support the translation of these findings into practice, the thematic and socioecological findings were translated into a consumer-informed socioecological framework for community falls prevention care (Figure 3). The framework highlights modifiable influences and potential opportunities to improve timely access to acceptable and affordable falls prevention care.

**Figure 3:**
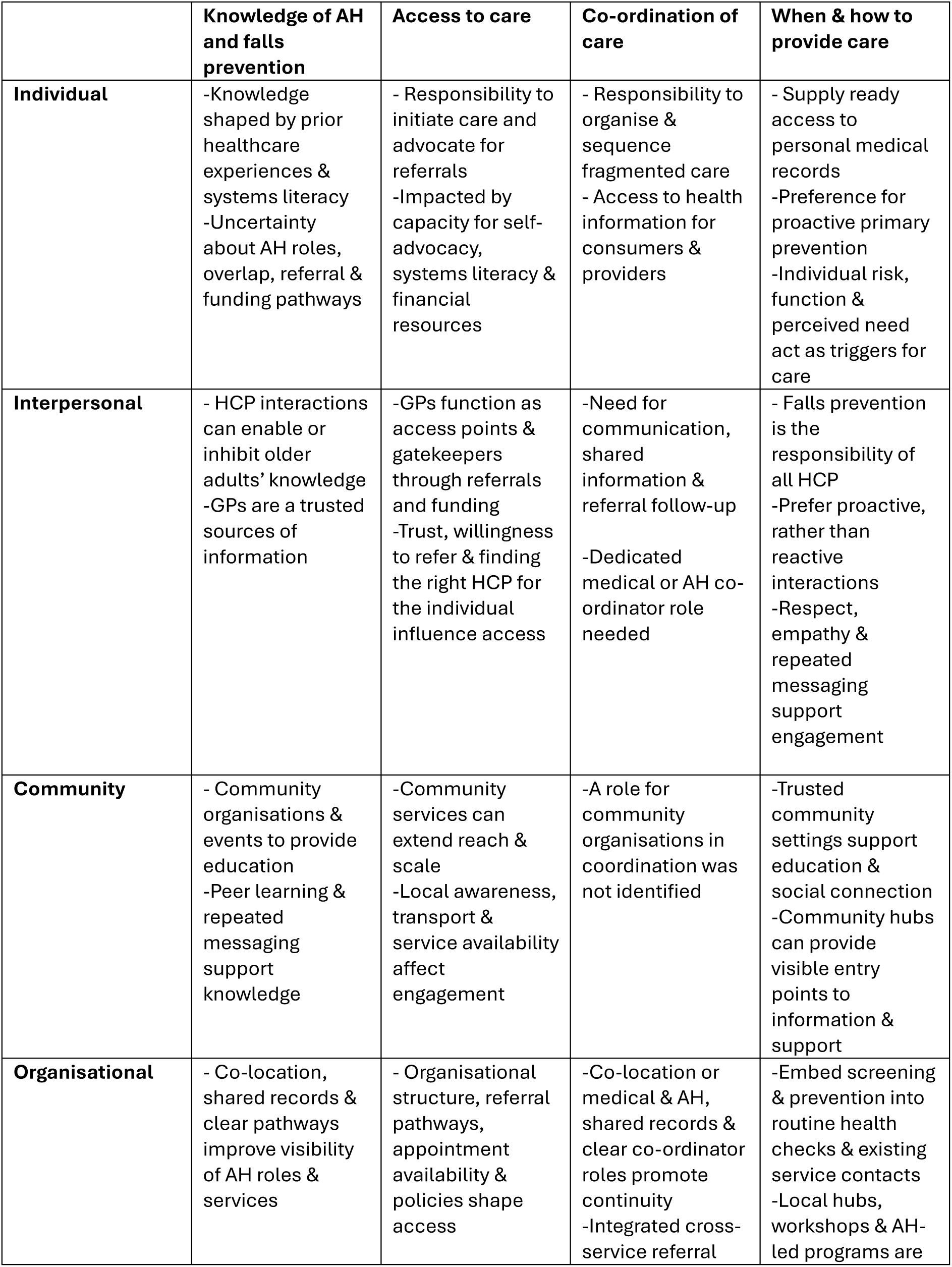
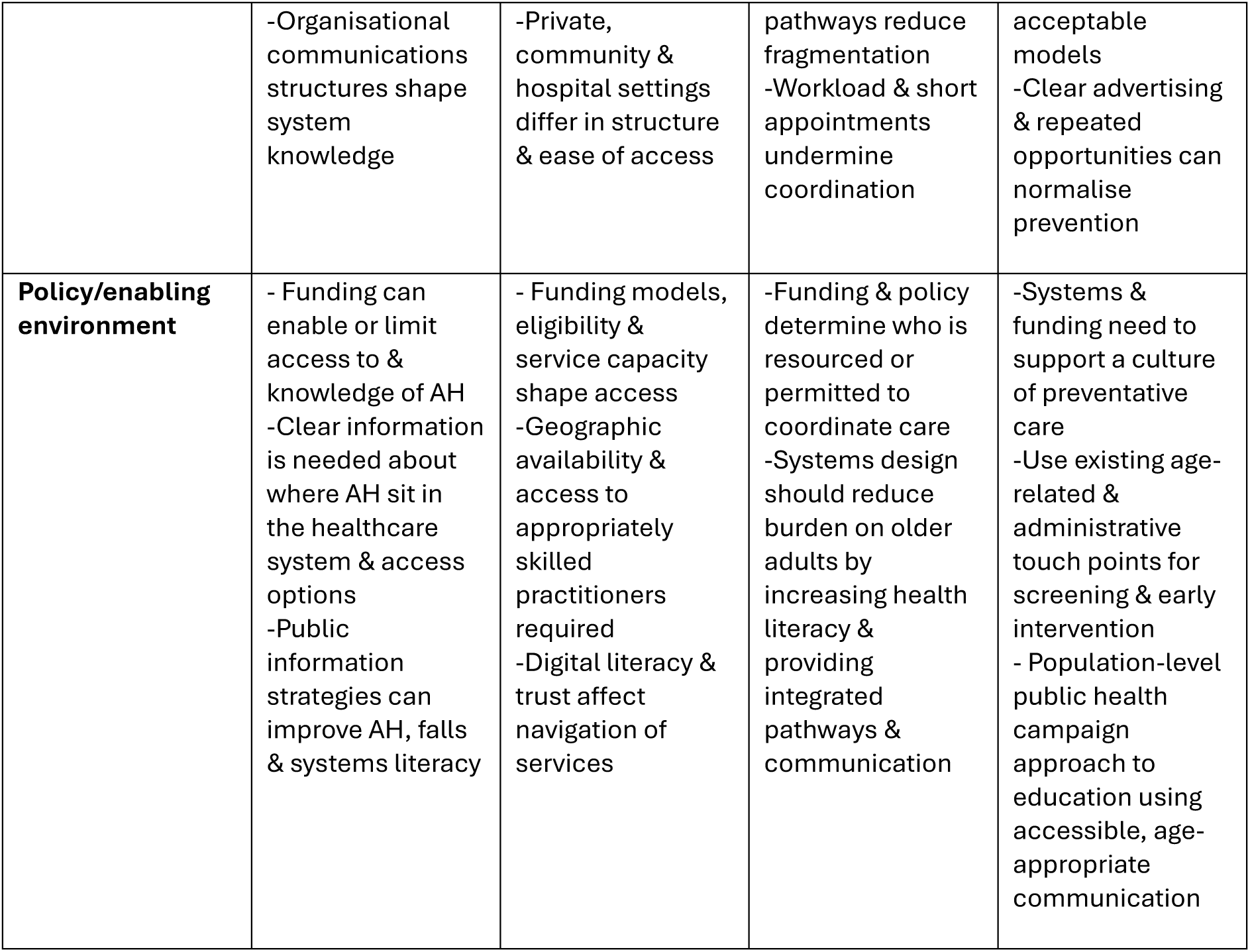
Consumer-informed socioecological framework for community interprofessional falls prevention care Abbreviations: AH, allied health; GP, general practitioner; HCP, healthcare professionals

## Strengths and limitations

This study should be interpreted in light of several limitations. As a qualitative focus group study conducted within a specific health system, findings may not be generalisable to all contexts and may reflect the experiences of older adults who w thoseere sufficiently engaged in healthcare to participate. Although both face to face and digital recruitment strategies were utilised, online recruitment may have influenced inclusion of older people with lower digital literacy. Recruitment across metropolitan and regional settings increased the diversity of perspectives represented. However, as the number of participants in each region was limited, the study does not propose to directly compare the groups but to explore how geographic region, amongst other contextual factors, influenced experiences of care.

Group dynamics and self-reported accounts may have influenced the breadth and depth of some participants’ perspectives and participation. Views of culturally and linguistically diverse populations, as well as Aboriginal and Torres Strait Islander Peoples should be explored to complement this study.

Key strengths of this study include its consumer-centred focus and the application of the socioecological framework enabling the interpretation of the perceptions across individual, interpersonal, community, organisational, and systems levels. The pragmatic orientation of the study design supported practice, service and policy-related translation of the data. Inclusion of participants both with and without a history of falls and falls prevention care, broadened the range of perspectives captured to include views developed through experience of care, as well as expectations of primary preventative care. Recruitment in both metropolitan and regional areas increased contextual diversity and supports the transferability of findings. Together these factors provide novel insight into older adults’ experiences and perceptions of interprofessional falls prevention care which have the potential to improve access to interprofessional collaborative care.

## Conclusion

This study highlights that falls prevention for community-dwelling older adults remains constrained by fragmented care and misalignment across individual, interpersonal, organisational, community and policy levels. Despite the participants strongly supporting preventative care, access to care is frequently reactive and consumer driven, and both enabled and constrained by policy and funding. A reliance upon individual relationships created variability in knowledge, access and coordination of care, further impacted by organisational structures and policies. These findings underscore a reciprocal process whereby knowledge enables access; access without co-ordination remains reactive; coordination enables timely, preventative care; and perspectives about when and how care should occur are shaped by features across all three themes. The current work highlights the need for whole-system reform that embeds falls prevention in routine healthcare for older adults, strengthens knowledge of AH roles and scope in the context of falls prevention, and aligns funding, coordination and public health strategies to deliver equitable and sustainable falls prevention care for older adults.

## Data availability

Data is not available due to planned additional analyses.

## Author contributions

A.L., B.V., R.L. and J.F. contributed to conceptualization and methodology. A.L., R.W., N.T. and B.V. contributed to the investigation. A.L. performed project administration, data curation, wrote the original draft and visualisation. All authors contributed to the writing review and editing. D.B., B.V., R.L. and J.F. contributed to supervision.

