## Supplementary materials for "Interprofessional falls prevention in community settings: Older adults’ perspectives through a socioecological lens"

**Supplementary material A: Focus group guide**

Welcome everyone and thank you for coming today. My name is (insert name) and I will be hosting this focus group. The purpose of this session is to gather your thoughts and experiences on falls prevention for older adults living in the community. The information you provide will help us to better understand the roles of the allied health professions and health professional collaboration and has the potential to improve falls prevention care and outcomes for older adults.

Before we begin, I would like to go over a few important points regarding your participation.

Your participation today is entirely voluntary, and you can withdraw from the focus group at any time. If there are topics you do not wish to discuss, you are free to not participate in those discussions. Please advise me of any issues throughout the session. To maintain the confidentiality of the other participants we ask that following the focus group, you do not discuss the contents with anyone else.

To ensure a productive discussion, please keep the following guidelines in mind:

- Please treat all members of the group with respect throughout the discussion.
- Please give everyone a chance to speak and listen carefully to what others are saying.
- To ensure everyone’s voice is recorded clearly, please avoid interrupting others.
- We want to stay focused on the subject at hand. If you have thoughts or ideas that aren’t directly related to the discussion, feel free to share them at the end
- We’ll be mindful of everyone’s time. Please be concise when sharing your thoughts to allow everyone time to contribute

In a moment I will commence the recording. Does anyone have any comments or questions before we begin?

The first group of questions I’m going to ask you, are about which healthcare professionals you’ve received falls prevention care from and who you would like to receive care from.

To begin with, please take a moment to review the list of healthcare professionals that I have provided you with. This is the same list that was sent to you when your focus group was confirmed.

(Give the group 2 minutes to review the list)

1. From the list of allied health professions I’ve given you, which do you think have a role in falls prevention and what is that role?
2. For those of you who have had a previous fall which healthcare professionals did you see for this?
   a. Which of them provided you with falls prevention advice to help avoid further falls?
   b. What was the advice?
   c. Did any of them refer you to other healthcare professionals to receive further care?
3. For those of you who have not had a previous fall, have you had any healthcare professionals provide you with falls prevention advice or care?
   a. If so, who and what advice or care did they provide
   b. Did any of them refer you to other healthcare professionals to receive further care?
4. What factors do you think should trigger an allied healthcare professional to assess your risk of falling?
   a. Examples if needed: balance problems, eyesight problems, being nervous about falling

The next series of questions relate to interprofessional care. Interprofessional care is when you receive care from two or more healthcare professionals for the same problem, in this case, falls prevention care.

Based on this definition of interprofessional care:

1. What are the barriers you face when trying to follow advice on seeing another healthcare professional when you are referred to them?
   a. Examples if needed: cost, location of different professional, understanding why you need to see each professional
2. What facilitates or helps you when you are trying to follow the advice to see another healthcare professional when you are referred to them?
   a. Examples if needed: understanding why you need to see each professional, communication between professionals

**Supplementary material B: List of allied health professions provided to focus group participants**

**Arts therapy**

Arts therapists use visual art-making, drama, dance and movement to improve physical, mental and emotional well-being. They work with individuals or groups using arts processes such as painting to create meaning, rather than focussing on the end product.

Arts therapy can be helpful for people who cannot verbalise their feelings due to developmental, cognitive or other conditions.

**Audiology**

Audiologists are experts in hearing loss and balance disorders. Audiologists can help people of all ages experiencing hearing loss with the use of hearing aids and other assistive technologies to improve their ability to communicate.

**Certified practicing nutritionist**

CPNs are bona fide primary care nutrition practitioners who practise Clinical Nutrition, which includes dietary modification (applied within a Clinical Nutrition/Nutritional Medicine paradigm) and the prescribing of dietary supplements for both nutrient repletion and complex nutritional medicine purposes.

**Chiropractic**

A chiropractor diagnoses and offers treatment for back pain and disorders of the musculoskeletal system. The treatment may include manipulation, massage or ergonomic advice. Chiropractors in Australia are a nationally registered and a regulated healthcare profession.

**Counsellors & psychotherapists**

Counsellors and Psychotherapists work with clients to benefit their mental health and wellbeing. They use a range of interventions including talking therapies as well as creative and experiential therapies (art, music, dance, eco and animal therapies) to build a positive therapeutic relationship which supports self-awareness and resolves identified concerns.

**Credentialled diabetes educators**

A Credentialled Diabetes Educator (CDE) is a health professional who is an expert in diabetes education and management. They support people in self-managing their diabetes, overcoming challenges, and leading their healthiest life possible.

**Dietetics**

Dietitians are experts in food and nutrition. They provide guidance about how to appropriately manage diets and nutrition for people who may be affected by health conditions such as diabetes, overweight and obesity, cancer, heart disease, renal disease, gastro-intestinal diseases and food allergies. A dietitian can help people maintain their health and reduce their risk of developing chronic disease.

**Exercise physiology**

Accredited exercise physiologists specialise in clinical exercise interventions for people with a broad range of health issues. Those people may be at risk of developing, or have existing, medical conditions and injuries. The aims of exercise physiology interventions are to prevent or manage acute, sub- acute or chronic disease or injury, and assist in restoring one’s optimal physical function, health or wellness. These interventions are exercise-based and include health and physical activity education, advice and support and lifestyle modification with a strong focus on achieving behavioural change.

**Genetic counselling**

Genetic counsellors have specialist knowledge in human genetics, counselling and health communication skills.  They work as part of a team, usually with medical specialists such as clinical geneticists, oncologists, obstetricians, neurologists and cardiologists.

A genetic counsellor provides information to individuals and families about genetic conditions. That may involve learning about how they are inherited or who in the family may be at risk of developing a particular condition. Genetic counsellors also provide emotional and practical support to help people adjust to living with, or being at risk for, a genetic condition.

**Medical radiations**

A Radiographer/Diagnostic Radiographer/Medical Imaging Technologist is responsible for producing high quality medical images that assist medical doctors and other practitioners to describe, diagnose, monitor and treat a patient’s injury or illness. As part of a diagnostic health team, radiographers are highly skilled individuals who operate extremely advanced technical equipment such as MRI scanners (magnetic resonance imaging), CT (computed tomography) and mobile X-ray machinery. Radiography uses both ionising and non-ionising radiation in the imaging process.

**Music therapy**

Music therapy is a research-based practice and profession in which music is used to actively support people as they strive to improve their health, functioning and well-being. Music therapists incorporate a range of music-making methods within and through a therapeutic relationship to address individual client goals.

**Occupational therapy**

Occupational therapists focus on promoting health and wellbeing by enabling people to participate in the everyday occupations of life, such as self-care activities including showering, dressing, preparing food; productive activities such as education, work, volunteering and caring for others; and leisure/social activities, such as being part of a community group, engaging in a hobby, and being part of a friendship group. Occupational therapists play a particularly crucial role in enabling people experiencing disability to identify and implement methods that support their participation in occupations. This may include modifying an activity or an environment.

**Optometry**

Optometrists are experts in eye health, trained to prescribe spectacles and contact lenses and treat a range of eye conditions such as dry eye, allergies and infections. Optometrists provide a wide range of services including vision testing, prescription of glasses and/or contact lenses, assessment and reporting on fitness to drive and vision problems in children such as strabismus and amblyopia.

**Orthoptics**

Orthoptists are healthcare professionals who are trained in the assessment, diagnosis and treatment and rehabilitation of patients with eye disorders. Orthoptists specialise in children’s vision, eye movement disorders and low vision care and rehabilitation.

**Orthotics/prosthetics**

Orthotists/prosthetists help people to reach their goals, such as increased community participation and movement. They do this through the provision of orthoses (splints and braces) and prostheses (artificial limbs) and associated clinical services. Orthotists/prosthetists provide devices to increase mobility and independence for people experiencing illness or disability.

**Osteopathy**

An osteopath has a clinical focus on the way the body works, in strains or injuries and in human movement. They provide direct manual therapy interventions including exercise prescription, needling, education and associated lifestyle advice to improve movement, reduce pain and manage and/or treat a range of physical impairments.

**Paramedic practitioners**

All paramedics can respond to emergencies. In addition to emergency healthcare skills, many Paramedics have additional primary care capabilities. These specialist paramedics can treat a wider range of health conditions. Paramedics with additional primary care skills are “generalists” and work alongside Doctors and Nurse Practitioners. They also work collaboratively and in team-based arrangements with other allied health professionals. With advanced capabilities, these specialist paramedics can assess, diagnose, and treat a broad range of patients. Paramedic Practitioner is the top tier of paramedics with additional primary care capabilities.

**Pedorthist custom makers**

A Pedorthist Custom Maker* is a health professional trained in the analysis and treatment of gait and foot & ankle problems. Pedorthists are experts in the use of both prefabricated and custom-made orthopaedic footwear, foot orthotics, and ankle braces, to provide practical and positive solutions to improve people’s daily life.

**Perfusion**

Perfusionists operate the heart-lung bypass machine (also known as cardiopulmonary bypass) during heart surgery to maintain a safe and stable condition while the heart is stopped for repair. Perfusionists may operate such equipment during any medical situation where it is necessary to support or temporarily replace the patient’s cardio-pulmonary or circulatory function.

**Pharmacy**

Pharmacists play an integral role ensuring patients receive high quality medicine management, providing essential services including: medicine reconciliation, clinical review, patient counselling and efficient supply. Pharmacists provide drug information and advice to health professionals and the community as well as contributing to and conducting clinical trials of new or improved treatments.

**Physiotherapy**

Physiotherapists are experts in the structure of the human body and its movement. They work with people of all ages to treat a broad range of health conditions including sports injuries and musculoskeletal conditions as well as chronic health conditions such as diabetes, obesity, osteoarthritis and stroke. Physiotherapists are involved in the assessment, diagnosis, planning and management of patient care.

**Podiatry**

A podiatrist is an expert in foot care. Podiatrists help people in the care of their lower limbs including the foot and ankle and may also be involved in supporting older people to reduce their risk of falling. The podiatrist’s scope of practice includes areas such as paediatrics, diabetes, sports injuries, structural problems, treatment of the elderly as well as general foot care.

**Psychology**

Psychologists are experts in human behaviour who can help people change the way they think, feel, behave and react. Psychologists study the brain, memory, learning and processes around human development. Psychological treatments can be used to help individuals, families, groups and organisations.

**Rehabilitation counselling**

Rehabilitation Counsellors facilitate social, educational and economic inclusion for people experiencing illness, injury, disability or disadvantage. Rehabilitation Counsellors play a critical role in the provision of assessment, case management, counselling and service provision to support people with disability, illness or injury to achieve their employment, volunteer or study goals. With advanced training in these specific life domains, Rehabilitation Counsellors are uniquely qualified in this respect.

**Social work**

Social workers support people to make change in their lives to improve their personal and social well-being. This happens by identifying issues that require change and connecting people with support such as secure housing or family therapy. Social workers have knowledge of human behaviour and development, life cycle stages, families and social networks, disability and health, including mental health.

**Sonography**

A sonographer performs specialised diagnostic examinations using high frequency ultrasound (sonography). People may require ultrasound scans for a range of conditions from pregnancy to more complex health conditions as prescribed by their health practitioner.

**Speech pathology**

Speech pathologists study, diagnose and treat communication disorders, including difficulties with speaking, listening, understanding language, reading, writing, social skills, stuttering and using voice. They work with people who have difficulty communicating because of developmental delays, stroke, brain injuries, learning disability, intellectual disability, cerebral palsy, dementia and hearing loss, as well as other problems that can affect speech and language. People who experience difficulties swallowing food and drink safely can also be helped by a speech pathologist.
